# The Global Parkinson’s Disease Genetics (GP2) Genome Browser

**DOI:** 10.64898/2025.12.29.25343143

**Authors:** Zih-Hua Fang, Riley H. Grant, Dan Vitale, Carlos F. Hernandez, Samantha Hong, Hampton L. Leonard, Mary B. Makarious, Lara M. Lange, Matthew Solomonson, Peter Heutink, Allison A. Dilliott, Kamalini Ghosh Galvelis, Mike A. Nalls, Andrew B. Singleton, Cornelis Blauwendraat, the Global Parkinson’s Genetics Program (GP2)

## Abstract

**Background:** Large-scale sequencing initiatives have generated extensive genomic resources essential for variant interpretation, yet their effective use often requires bioinformatics expertise. To support identification of Parkinson’s disease (PD) risk and disease-causing variants, we developed an open-access, summary-level genomic data browser.

**Methods:** We performed uniform joint variant calling to harmonize whole-genome sequencing (WGS) data from AMP-PD Release 4, GP2 Data Releases, and additional controls from the Alzheimer’s Disease Sequencing Project. Clinical exome sequencing (CES) data from GP2 Release 8 was also included.

**Results:** The integrated dataset includes 31,665 WGS and 9,559 CES samples, spanning eleven ancestries and over 300 million variants.

**Conclusion:** The GP2 Genome Browser is a lightweight, flexible platform providing intuitive gene- and variant-level summaries with ancestry-stratified allele frequencies and functional annotations. It is open source and freely accessible at https://gp2.broadinstitute.org, enabling broad access to PD genomic data and supporting global research efforts.

## Introduction

Precise molecular mechanisms underlying Parkinson’s Disease (PD) remain largely uncertain. The known risk factors include aging, environmental exposures, and complex genetic factors. Advances in genetic research have identified both rare causative and common risk variants, primarily in individuals of European ancestry^1^. Extending the data to large, ancestrally diverse cohorts will improve our understanding of differences in genetic susceptibility to PD across populations, which is crucial for developing efficient therapies that target patients with diverse ancestries.

Whole-genome sequencing provides a more comprehensive view of the genetic architecture by capturing a broad range of genetic variation, including rare variants that may have a greater phenotypic impact. However, analyzing and interpreting these large-scale data requires substantial bioinformatics expertise, underscoring the importance of continued data aggregation and harmonization to improve variant interpretation^2^. A prior initiative, the Parkinson’s Disease DNA Variant Browser^3^, primarily featured data from individuals of European ancestry. Building on this foundation, we leveraged whole-genome (WGS) and clinical-exome (CES) sequencing data available from the Global Parkinson’s Genetics Program (GP2) Data Releases, Accelerating Medicines Partnership - Parkinson Disease (AMP-PD, https://amp-pd.org/) and additional controls from the Alzheimer’s Disease Sequencing Project^4^. Using these datasets, we developed the GP2 Genome Browser, which integrates and displays variant- and gene-level data across diverse genetic ancestries.

## Methods

### Whole-genome sequencing (WGS) data

We harmonized the WGS data available from AMP-PD release 4, GP2 Data Release 10 (DOI: 10.5281/zenodo.15748014)^5^ and additional 4,278 controls of European ancestry available from the Alzheimer’s Disease Sequencing Project (DOI: 10.60859/z6z9-9692) ^4^ by generating single-sample variant calls from the alignment files with DeepVariant v.1.6.1^6^ (https://github.com/google/deepvariant) followed by joint genotyping of single nucleotide variants (SNVs) and short indels using GLnexus v1.4.3 (https://github.com/dnanexus-rnd/GLnexus) with the preset DeepVariant WGS configuration^7^. We set genotypes to be missing after genotype quality control, defined as genotype quality >= 10, read depth >= 5, and heterozygous allele balance between 0.2 and 0.8, and retained high-quality variants with a call rate >= 0.95. After sample quality control using the quality metrics defined by AMP-PD^8^, we retained 31,665 samples for downstream analyses. We used KING v.2.3.0 (https://www.kingrelatedness.com, RRID:SCR_009251)^9^ to infer relatedness up to the second-degree relatives to confirm known relationships and identify cryptic familial relationships. Genetic ancestry was determined using GenoTools v1.2.3 (https://github.com/GP2code/GenoTools) with the default settings^10^. Variant annotation was performed with Ensembl Variant Effect Predictor v111 (http://www.ensembl.org/info/docs/tools/vep/index.html, RRID:SCR_007931)^11^. Intergenic variants were excluded from the browser display. Allele frequencies for the remaining variants were calculated and stratified by phenotype and genetic ancestry. PD patients were defined as individuals diagnosed with PD, excluding those enrolled via targeted recruitment of known mutation carriers (such as *LRRK2* and *GBA1*). Controls were defined as healthy individuals, excluding those recruited as known mutation carriers, individuals recruited through family-based designs, and those with a positive family history of PD. Related individuals were retained because the dataset was generated for association studies using mixed logistic regression models, with the results planned for integration into the browser at a later stage. The remaining individuals were grouped together as ‘Other’ phenotypes for allele frequency calculation. This ‘Other’ phenotypes group contains other primary phenotypes, including but not limited to prodromal stages of PD (e.g., hyposmia and REM sleep behavior disorder), atypical parkinsonism, other neurodegenerative or movement disorders, and dementia; for further details, see the GP2 release notes (https://gp2.org/updates/?post_type=Data%20Release#results).

### Clinical-exome sequencing

We included 10,454 samples with clinical-exome data available from PD GENEration^12^ as part of GP2’s Data Release 8 (DOI 10.5281/zenodo.13755496)^13^. The sequence data processing followed the same pipeline as the WGS data mentioned above. We performed joint-genotyping using GLnexus (v1.4.3) with the preset DeepVariant WES configuration and applied the same criteria for genotype, sample, and variant quality control. After excluding CES samples identified as genetic duplicates of individuals in the WGS dataset, a final set of 9,559 samples was retained for downstream frequency calculation.

## Results

We performed uniform joint variant calling to harmonize WGS data from AMP-PD Release 4, GP2 Data Release 10^5^, and additional controls of European ancestry available from the Alzheimer’s Disease Sequencing Project^4^. Additionally, we included the CES data from GP2 Data Release 8^13^ for calculating allele frequency. After excluding samples that failed quality control or were duplicated, and removing variants that failed quality control or were located in intergenic regions, we retained 303,505,959 variants from 31,665 individuals with WGS data and 1,499,072 variants from 9,559 individuals with CES data. The WGS cohort comprised individuals of European (71.4%), East Asian (8.0%), Ashkenazi Jewish (6.1%), African (5.3%), and other ancestries, while the CES cohort was predominantly European (82.2%) (Table 1).

**Table 1.** Number of participants by genetically determined ancestry included in the whole-genome sequencing (WGS) and clinical exome sequencing (CES) datasets.

|  | WGS |  |  | CES |
| --- | --- | --- | --- | --- |
| Ancestry | Case | Control | Other | Case |
| African Admixed | 141 | 126 | 14 | 176 |
| African | 809 | 837 | 19 | 77 |
| Ashkenazi Jewish | 491 | 323 | 1123 | 0 |
| Complex Admixture | 110 | 37 | 37 | 278 |
| Central Asian | 338 | 327 | 147 | 48 |
| East Asian | 1848 | 344 | 343 | 130 |
| Finnish European | 25 | 7 | 6 | 282 |
| Latino and Indigenous people of the Americas | 287 | 29 | 42 | 564 |
| Middle Eastern | 478 | 308 | 47 | 52 |
| Non-Finnish European | 8252 | 7559 | 6804 | 7860 |
| South Asian | 284 | 15 | 108 | 92 |
| Total | 13063 | 9912 | 8690 | 9559 |

\*Cases were defined as individuals diagnosed with PD, excluding those recruited based on genetic status. Controls were defined as healthy individuals, excluding those recruited based on genetic status or through family-based recruitment. The 'Other' phenotypes group contains other primary phenotypes, including but not limited to prodromal stages of PD (e.g., hyposmia and REM sleep behavior disorder), atypical parkinsonism, other neurodegenerative or movement disorders, and dementia; for further details, see the GP2 release notes (https://gp2.org/updates/?post_type=Data%20Release#results).

The browser adopts a gene page interface similar to that of the Genome Aggregation Database (gnomAD, http://gnomad.broadinstitute.org, RRID:SCR_014964)^14^. The front page includes a search bar with autocomplete functionality allowing users to search gene symbols and Ensembl gene IDs. The gene page provides an overview of gene-level information (Figure 1A). It integrates reference data from external resources, including gnomAD and other large-scale case-control studies of neurological phenotypes, such as the Bipolar Exome (BipEx) sequencing project^15^ and the Epi25 Collaborative^16^, among others (Figure 1B). An exon summary plot illustrates the positions of detected variants within the selected ancestry group and the total number of variants stratified by case and control status, with color coding representing their functional annotations (Figure 1C). Variants detected within the selected gene are also displayed on this page, stratified by genetic ancestry (Figure 1D). The variant page presents a set of annotations for each variant, including the CADD Phred score^17^, ClinVar classification^18^, and dbSNP rsID^19^, along with a table summarizing population frequencies across genetic ancestries and datasets (Figure 2).

**Figure 1.**
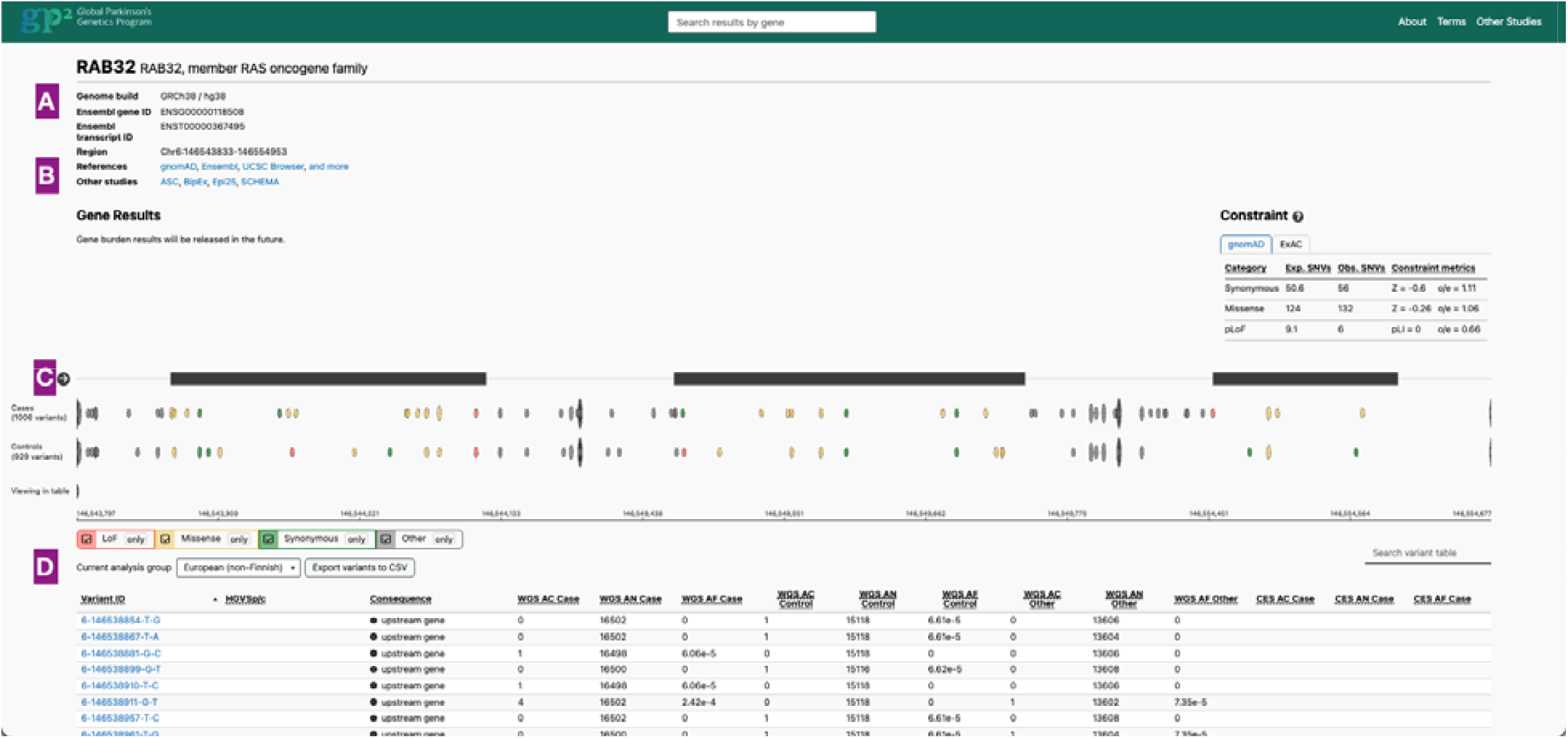
Gene page. (A) Gene information (B) Links to various external resources, as well as constraint information available from gnomAD 4.1 (C) The position of the variants in the selected genetic ancestry stratified by case and control (D) A table of all variants is provided with additional annotation information and links to variant pages.

**Figure 2.**
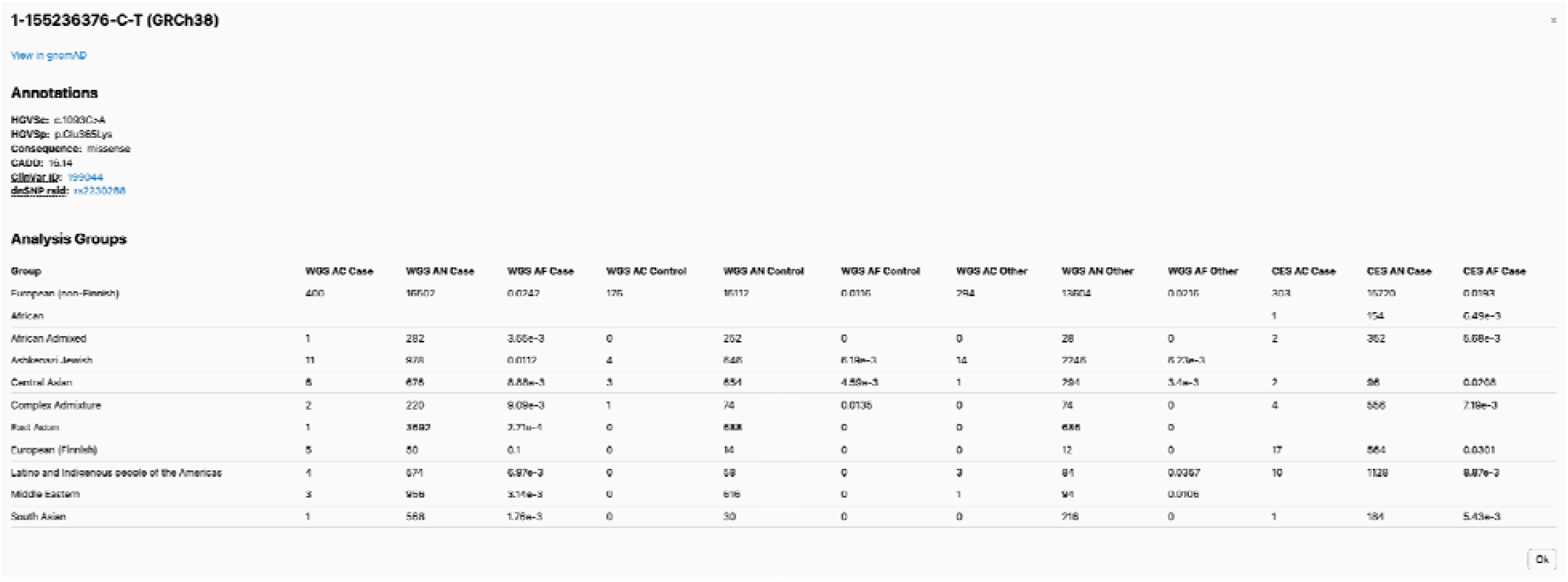
Variant page. A set of variant annotations is displayed, including links to dbSNP and ClinVar. Allele frequency information is displayed for each genetic ancestry and phenotypes.

We examined known pathogenic and risk variants in established PD genes to validate their distribution across genetic ancestries. In the *LRRK2* gene, the p.G2019S variant— one of the most common pathogenic variants—was detected across several ancestries but was absent in African admixed, Central Asian, East Asian, South Asian, and Finnish European populations. The highest allele frequency among PD patients (0.04) was observed in individuals of Ashkenazi Jewish ancestry, followed by Middle Eastern ancestry (0.01) in the WGS dataset. These findings are consistent with previous reports showing that *LRRK2* p.G2019S is particularly common in Ashkenazi Jewish and North African Berber Arab populations^20,21^. Conversely, the East Asian–specific risk variant p.G2385R in *LRRK2* was identified in non-Finnish European, Central Asian, and East Asian populations, with a markedly higher allele frequency in East Asian PD patients (0.05) compared to non-Finnish European PD patients (1.21×10□□) in the WGS dataset, also consistent with prior findings^22^. Additionally, the recently identified *RAB32* p.S71R variant^23,24^ was detected in non-Finnish European and Middle Eastern populations but was absent in all other ancestries. Taken together, these findings demonstrate that the browser can facilitate the investigation of both shared and ancestry-specific patterns in the prevalence and spectrum of SNVs and indels relevant to PD research.

## Discussion

We present the GP2 Genome Browser (https://gp2.broadinstitute.org), a public platform that allows for rapid querying of specific genes and variants within the largest and most diverse sequencing collection assembled for PD research (13,063 PD vs 9,912 Others vs 8,690 Control). Upon input of a gene HGNC symbol or Ensembl gene ID, the browser presents the variants found within that gene, categorized by functional consequence with the frequencies stratified across genetic ancestries and phenotypes. For a specific variant, the browser displays several annotations, including functional consequence, allele frequency across genetic ancestries and phenotypes, ClinVar information and dbSNP rsID. These functionalities enable users to retrieve the frequency of a variant of interest (identified in a PD patient or within a family) and compare shared versus ancestry-specific patterns across populations.

The current version of the GP2 Genome Browser has several limitations. First, it currently displays data only from the variants located within upstream and downstream 5Kb of the gene body. While these regions are often the most relevant for identifying disease-causing variants, many non-coding regions also play critical roles in human disease and remain active areas of research^25^. Second, the browser does not currently display zygosity distributions, which are relevant for investigating variants that follow a recessive mode of inheritance. Third, the frequencies for pseudogene-related variants in *GBA1*, including p.A495P, p.L483P, p.D448H, c.1263del, RecNciI, RecTL, and c.1263del+RecTL, might be underestimated when using the standard variant calling pipeline compared to the *GBA1* target caller, Gauchian^26^. Furthermore, some *GBA1* variants may be missed due to false negative calls. Finally, allele frequency estimates may be inflated due to the presence of related individuals in the datasets. Note that the browser is intended to display results from association analyses that use mixed logistic regression models, along with the corresponding datasets used for those analyses.

In summary, we present an online resource designed for the PD research community to efficiently access annotated genomic information on genes and variants through a user-friendly interface, similar to gnomAD—not requiring coding or data science expertise required. Researchers can use the browser to explore variant frequencies between PD patients, PD-related phenotypes, and controls, across genetic ancestries. The browser’s data can also be used to complement other researchers’ own analyses with data from large-scale genomic datasets. We plan to update the browser biannually with future GP2 releases, refine its features, and incorporate association results for both single variants and gene-based burden analyses as they become available.

## Data Availability

GP2 partnered with the online cloud computing platform Accelerating Medicines Partnership - Parkinson's Disease (AMP-PD; https://amp-pd.org) to share data generated by GP2. Qualified researchers are encouraged to apply for direct access to the data through AMP-PD. The GP2 and AMP-PD datasets analysed during the current study are available through AMP-PD (https://amp-pd.org).
Data used in the preparation of this article were obtained from the Global Parkinson's Genetics Program (GP2; https://gp2.org). Specifically, we used Tier 2 data from GP2 release 8 and 10 (10.5281/zenodo.13755496, Release 8; 10.5281/zenodo.15748014, Release 10). GP2 data can be accessed through AMP-PD (https://amp-pd.org).
All code generated for this article, and the identifiers for all software programs and packages used, are available on GitHub (https://github.com/GP2code/GP2-Genome-Browser) and were given a persistent identifier via Zenodo (DOI 10.5281/zenodo.17903069).

## Acknowledgment

Clinical-exome sequencing data (DOI 10.5281/zenodo.13755496, Release 8) and whole-genome sequencing data (DOI 10.5281/zenodo.15748014, Release 10) used in the preparation of this article were obtained from the Global Parkinson’s Genetics Program (GP2) and AMP-PD.

This project was supported by the Global Parkinson’s Genetics Program (GP2; https://gp2.org). GP2 is funded by the Aligning Science Across Parkinson’s (ASAP) initiative and implemented by The Michael J. Fox Foundation for Parkinson’s Research (MJFF). For a complete list of GP2 members see https://doi.org/10.5281/zenodo.7904831.

The AMP® PD program is a public-private partnership managed by the Foundation for the National Institutes of Health and funded by the National Institute of Neurological Disorders and Stroke (NINDS) in partnership with the Aligning Science Across Parkinson’s (ASAP) initiative; Celgene Corporation, a subsidiary of Bristol-Myers Squibb Company; GlaxoSmithKline plc (GSK); The Michael J. Fox Foundation for Parkinson’s Research; Pfizer Inc.; AbbVie Inc.; Sanofi US Services Inc.; and Verily Life Sciences. ACCELERATING MEDICINES PARTNERSHIP and AMP are registered service marks of the U.S. Department of Health and Human Services. Clinical data used in preparation of this article were obtained from the MJFF-sponsored LRRK2 Cohort Consortium (LCC). For up-to-date information on the study, visit www.michaeljfox.org./lcc. The LRRK2 Cohort Consortium is coordinated and funded by The Michael J. Fox Foundation for Parkinson’s Research. The investigators within the LCC provided data, but did not participate in the analysis or writing of this report. The full list of LCC investigators can be found at www.michaeljfox.org/lccinvestigators. PD GENEration is a study funded by the Parkinson’s Foundation and supported by GP2, a program of Aligning Science Across Parkinson’s (ASAP). Data from Alzheimer’s Disease

Sequencing Project (NG00067, DOI: 10.60859/z6z9-9692) used in this study were prepared, archived, and distributed by the National Institute on Aging Alzheimer’s Disease Data Storage Site (NIAGADS) at the University of Pennsylvania (U24-AG041689), funded by the National Institute on Aging. This research was supported by the Intramural Research Program, National Institute on Aging, National Institutes of Health, Department of Health and Human Services, project ZO1 AG000949. This work utilized the computational resources of the NIH STRIDES Initiative (https://cloud.nih.gov) through the Other Transaction agreement - Azure: OT2OD032100, Google Cloud Platform: OT2OD027060, Amazon Web Services: OT2OD027852. This work utilized the computational resources of the NIH HPC Biowulf cluster (https://hpc.nih.gov). The contributions of the NIH author(s) are considered Works of the United States Government. The findings and conclusions presented in this paper are those of the author(s) and do not necessarily reflect the views of the NIH or the U.S. Department of Health and Human Services.

## Data and Code Availability

GP2 partnered with the online cloud computing platform Accelerating Medicines Partnership - Parkinson’s Disease (AMP-PD; https://amp-pd.org) to share data generated by GP2. Qualified researchers are encouraged to apply for direct access to the data through AMP-PD. The GP2 and AMP-PD datasets analysed during the current study are available through AMP-PD (https://amp-pd.org).

Data used in the preparation of this article were obtained from the Global Parkinson’s Genetics Program (GP2; https://gp2.org). Specifically, we used Tier 2 data from GP2 release 8 and 10 (10.5281/zenodo.13755496, Release 8; 10.5281/zenodo.15748014, Release 10). GP2 data can be accessed through AMP-PD (https://amp-pd.org).

All code generated for this article, and the identifiers for all software programs and packages used, are available on GitHub (https://github.com/GP2code/GP2-Genome-Browser) and were given a persistent identifier via Zenodo (DOI 10.5281/zenodo.17903069).

## Authors’ roles

C.B. and Z.-H.F. conceived and designed the study. Z.-H.F., R.H.G., D.V., S.H., C.F.H., H.L.L., A.A.D., and K.G.G. contributed data, performed analyses, and developed the browser. M.B.M., L.M.L., M.S., P.H., M.A.N., P.H., A.B.S., and C.B. provided feedback on the browser design. Z.-H.F. wrote the first draft of the manuscript, and all authors contributed to editing the final version.

